# Vicarious traumatization in the general public, members, and non-members of medical teams aiding in COVID-19 control

**DOI:** 10.1101/2020.02.29.20029322

**Authors:** Zhenyu Li, Jingwu Ge, Meiling Yang, Jianping Feng, Mei Qiao, Riyue Jiang, Jiangjiang Bi, Gaofeng Zhan, Xiaolin Xu, Long Wang, Qin Zhou, Chenliang Zhou, Yinbing Pan, Shijiang Liu, Haiwei Zhang, Jianjun Yang, Bin Zhu, Yimin Hu, Kenji Hashimoto, Yan Jia, Haofei Wang, Rong Wang, Cunming Liu, Chun Yang

**Affiliations:** Department of Anesthesiology and Perioperative Medicine, The First Affiliated Hospital of Nanjing Medical University, Nanjing 210029, China; Department of Ultrasound Imaging, Renmin Hospital of Wuhan University, Wuhan 430060, China; Department of Anesthesiology, Tongji Hospital, Tongji Medical College, Wuhan 430030, China; Department of Anesthesiology, Renmin Hospital of Wuhan University, Wuhan 430060, China; Department of Critical Care Medicine, Renmin Hospital of Wuhan University, Wuhan 430060, China; Department of Anesthesiology, The First Affiliated Hospital of Zhengzhou University, Zhengzhou 450052, China; Department of Critical Care Medicine, The Third Affiliated Hospital of Soochow University, China; Department of Anesthesiology, The Second Affiliated Changzhou People’s Hospital of Nanjing Medical University, Changzhou 213000, China; Division of Clinical Neuroscience, Chiba University Center for Forensic Mental Health, Chiba 260-8670, Japan; Department of Orthopedics, The First Affiliated Hospital of Nanjing Medical University, Nanjing 210029, China; Department of Psychology, The First Affiliated Hospital of Nanjing Medical University, Nanjing 210029, China; Department of Nursing, The First Affiliated Hospital of Nanjing Medical University, Nanjing 210029, China

**Keywords:** COVID-19, vicarious traumatization, front-line nurses, non-front-line nurses, general public

## Abstract

Since December 2019, more than 79,000 people have been diagnosed with infection of the Corona Virus Disease 2019 (COVID-19). A large number of medical staff were dispersed for Wuhan city and Hubei province to aid COVID-19 control. Psychological stress, especially vicarious traumatization (VT) caused by the COVID-19 pandemic, should not be ignored. To address this concern, the study employed a total of 214 general public (GP) and 526 nurses to evaluate VT scores via a mobile app-based questionnaire. Results showed that the VT scores slightly increased across periods of aiding COVID-19 control, although no statistical difference was noted (*P* = 0.083). However, the study found lower scores for VT in nurses [median = 69; interquartile range (IQR) = 56–85] than those of the GP (median = 75.5; IQR = 62–88.3) (*P* = 0.017). In addition, the VT scores for front-line nurses (FLNs; median = 64; IQR = 52–75), including scores for physiological and psychological responses, were significantly lower than those of non-front-line nurses (nFLNs; median = 75.5; IQR = 63–92) (*P* < 0.001). Interestingly, the VT scores of the GP were significantly higher than those of the FLNs (*P* < 0.001). However, no statistical difference was observed compared with those of nFLNs (*P* > 0.05). Importantly, nFLNs are more likely to suffer from VT, which might be related to two factors, namely, gender [odds ratio (OR) = 3.1717; 95% confidence interval (CI) = 4.247–18.808; *P* = 0.002] and fertility [OR = 2.072; 95%CI = 0.626–24.533; *P* = 0.039]. Therefore, increased attention should be paid to the psychological problems of the medical staff, especially nFLNs, and GP under the situation of the spread and control of COVID-19. Early strategies that aim to prevent and treat VT in medical staff and GP are extremely necessary.

## Introduction

Since December 2019, the outbreak of COVID-19 in Wuhan has infected more than 70,000 individuals. China has taken active and effective actions to provide medical support for aiding in the control of the rapid spread of COVID-19. From January 24, 2020 (Chinese New Year’s Eve), China has sent more than 30,000 medical staff to Wuhan city and Hubei province to provide medical support. Researchers have validated that these efficient and feasible strategies and measures are timely and effective. Medical staffs often have a variety of psychological problems under a high-pressure and risk anti-pandemic situation.^1^ Therefore, psychological assessment and intervention in victims and rescuers, such as medical staff and volunteers, are of great importance for the control of large-scale disasters and pandemics. This notion is not only beneficial for early actions and measures for psychological intervention, but also for tremendously improving disaster and pandemic control and rapid social recovery.^2^

In 1996, Saakvitne and Pearlman first proposed vicarious traumatization (VT).^3^ The term initially referred to the phenomenon where professional psychotherapists are involuntarily affected by the bidirectional interactions of the relationship between consultation and interview due to long-term contact with patients with mental diseases. In other words, psychotherapists experienced mental symptoms similar to psychological trauma.^4^ Currently, the scope of application of VT is extended to a large number of cruel and destructive disasters, where the degree of damage exceeds psychological and emotional tolerance and indirectly leads to various psychological abnormalities.^5^ These psychological abnormalities are derived from sympathy for survivors of a trauma, which causes serious physical and mental distress, even mental breakdown.^6^

The main symptoms of VT such as anorexia, fatigue, physical decline, sleep disorder, irritability, inattention, numbness, fear, and despair are well recognized to be experienced by all individuals. Frequently, these symptoms are accompanied by trauma responses and interpersonal conflicts that even compel others to commit suicide.^7^ In this regard, the study proposes that medical staff, volunteers, and the GP will more or less experience VT during the spread and control of the COVID-19 pandemic. Therefore, identifying and providing intervention for VT at an early stage is important.

In this study, 214 GP and 526 nurses (i.e., 234 front-line nurses (FLNs) and 292 non-front-line nurses (nFLNs)), were employed and evaluated via the Chinese version of the VT evaluation scale. In addition, risk factors finally leading to VT among medical staff were evaluated. Therefore, our findings can likely provide theoretical basis and viable strategies for early psychological interventions during COVID-19 control.

## Methods

### Settings and participants

The study is descriptive in nature, utilizes a mobile phone app-based questionnaire survey, and was carried out during the COVID-19 pandemic from February 17, 2020 to Feb 21, 2020 (i.e., five weekdays). The Ethics Committee of the First Affiliated Hospital of Nanjing Medical University approved the study (approval number: 2020-SR-101). The study employed licensed registered nurses who worked in hospitals and GP (non-medical staff). Owing to the fact that the investigation was conducted during the COVID-19 pandemic, the current isolation policy calls for reduced face-to-face contact and communication and avoidance of large gatherings and activities. Therefore, an anonymous questionnaire was structured using a mobile app called “Sojump” (www.sojump.com) and pushed to individuals via WeChat after obtaining informed consent. Finally, a total of 740 individuals (i.e., 526 nurses and 214 GP), filled in the questionnaire.

### Demographic data and VT questionnaire

Demographic data included gender, age, hospital classification, years of working, departments, professional titles, undertaking management work or not, educational background, marriage status, single child or not, and fertile or not. The Chinese version of the VT questionnaire was compiled based on qualitative interviews with rescuers in the Wenchuan earthquake in China and existing international trauma-related scales, such as the Traumatic Stress Institute Belief Scale, Impact of Event Scale, and Vicarious Trauma Scale.^8-10^ The VT questionnaire adopted in the current study has a total of 38 items, which are composed of two dimensions, namely, physiological responses (11 items) and psychological responses [i.e., emotional responses (nine items), behavioral responses (seven items), cognitive responses (five items), and life belief (six items)]. Each question score ranged from 0 (never) to 5 (always). The higher the score, the more serious the VT. Cranach’s alpha for the questionnaire reached 0.93, whereas that for each dimension ranged from 0.73 to 0.92.

The cumulative variance contribution rate reached 52.56%, which indicates positive reliability and validity.

### Statistical analysis

The study presented continuous and normally distributed variables as means and standard error of mean. The independent sample t-test was used to assess group differences. Abnormally distributed data were described using the median and interquartile range (IQR: 25%–75%), whereas the Mann–Whitney U-test or Kruskal–Wallis H-test was used to assess group differences. Descriptive statistics involved frequencies (%) for categorical variables, and the chi-square test or Fisher’s exact test was used to assess group differences. The independent sample t-test, Mann–Whitney U-test, chi-square test, or Fisher’s exact test was adopted to compare variables between FLNs and nFLNs. Moreover, multiple linear regression analysis was used to determine the risk factors associated with the VT of nFLNs. Results were presented as OR and 95% CIs. Data were considered statistically significant when *P* < 0.05. Analyses were performed using SPSS version 20.0 (IBM Co. LTD, Chicago, IL, USA).

## Results

### VT score in FLNs

The study enrolled 526 license-registered nurses. Out of the 234 FLNs aiding in COVID-19 control, scores for VT, physiological responses, and psychological responses reached 64 (IQR = 52–75), 17 (IQR = 12–21), and 46.5 (IQR = 38–55), respectively. In addition, scores for physiological responses include scores for behavioral responses [13 (IQR = 10–15)], emotional responses [15 (IQR = 12–18.3)], cognitive responses [7 (IQR = 5–9)], and life beliefs [11 (IQR = 9–13)] (**Table 1**). Interestingly, the VT scores for FLNs across periods of aiding in COVID-19 control indicated a slight increase, although no statistical difference was observed (*P* = 0.083) (**Figure 1**).

**Table 1.** Severity of vicarious traumatization in 234 FLNs.

|  | Range of scores | Median (IQR) |  |
| --- | --- | --- | --- |
|  |  | Average scores of items | Scores |
| Vicarious traumatization | 38~126 | 1.7 (1.4-2.0) | 64 (52-75) |
| Physiological responses | 11~38 | 1.6 (1.1-1.9) | 17 (12-21) |
| Psychological responses | 27~88 | 1.7 (1.4-2.0) | 46.5 (38-55) |
| Behavioral responses | 7~24 | 1.9 (1.4-2.1) | 13 (10-15) |
| Emotional responses | 9~35 | 1.7 (1.3-2.0) | 15 (12-18.3) |
| Cognitive responses | 5~17 | 1.4 (1-1.8) | 7 (5-9) |
| Life beliefs | 6~24 | 1.8 (1.5-2.2) | 11 (9-13) |

Abbreviations: FLNs, first-line nurses; IQR, interquartile range.

**Figure 1.**
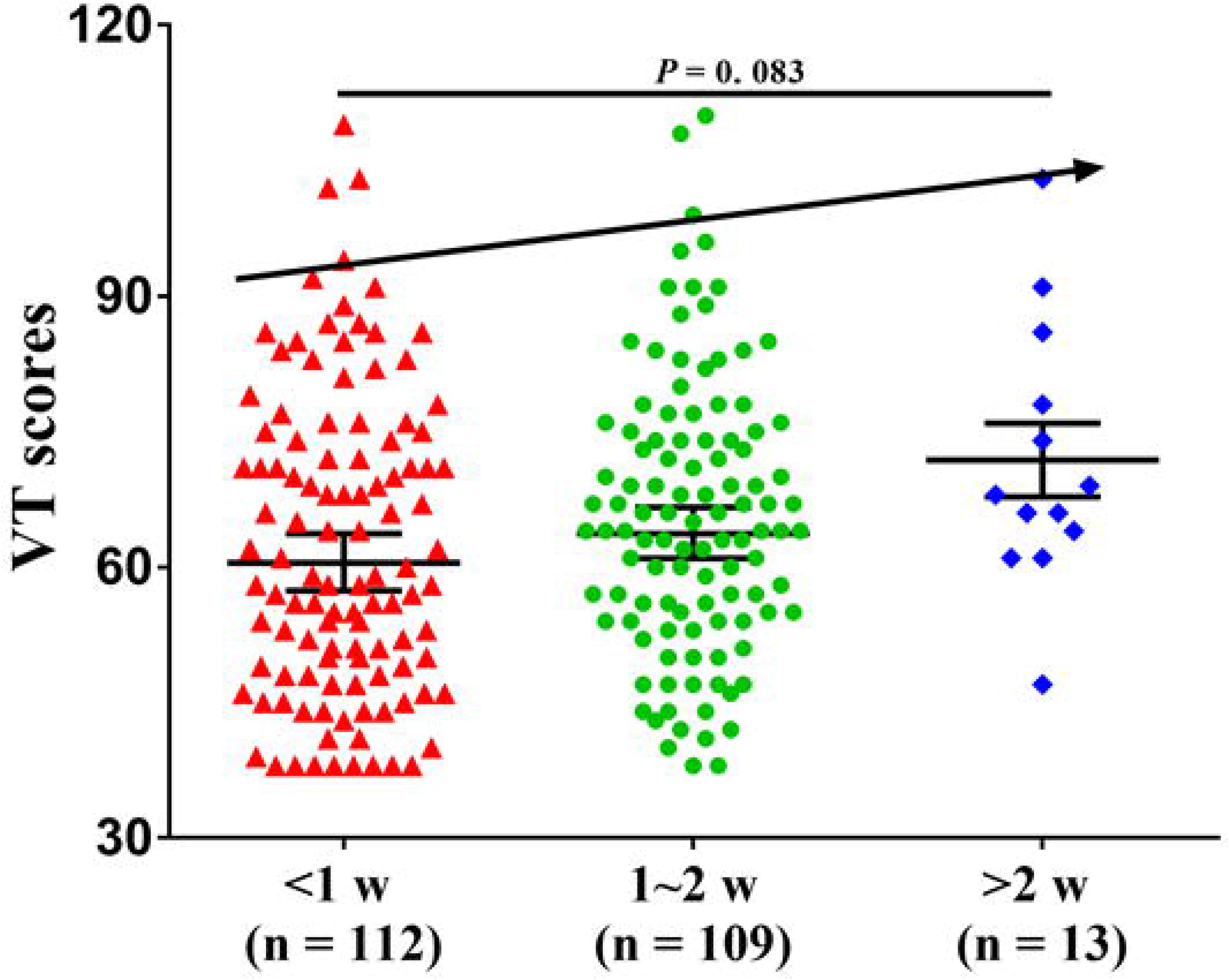
VT scores for FLNs across periods of aiding in COVID-19 control; VT scores (*P* > 0.05); FLNs, front-line nurses; VT, vicarious traumatization

### Comparison of general characteristic and VT scores between GP and nurses

A total of 740 individuals (i.e., 214 GP and 526 nurses) were enrolled in the study. Gender, age, marriage status, and fertility pointed to a significantly statistical difference between GP and nurses (**Table 2**). Importantly, VT scores for the GP [75.5 (IQR = 62–88.3)] demonstrated a significant increase compared with those of nurses [69 (IQR = 56–85)] (*P* = 0.017). The VT scores consist of scores for physiological responses and psychological responses. The scores for physiological responses failed to show a significant difference between GP [18 (IQR = 13–24)] and nurses [18 (IQR = 13–23)] (*P* = 0.656). In contrast, the scores for psychological responses were significantly higher in the GP [57 (IQR = 47–65.3)] compared with those in nurses [52 (IQR = 42–62)] (*P* = 0.003). The sub-items for psychological responses include behavioral responses [GP: 15 (IQR = 12–18), nurses: 14 (IQR = 11–17); *P* = 0.006], emotional responses [GP: 19 (IQR = 15–23), nurses: 17 (IQR = 13–21); *P* = 0.004], and life beliefs [GP: 13.5 (IQR = 11–17), nurses: 12 (IQR = 10–15); *P* = 0.004], but not cognitive responses [GP: 8 (IQR = 6–10), nurses: 8 (IQR = 6–10); *P* = 0.621], which demonstrated a significant increase in scores for GP than those for nurses (**Table 3**).

**Table 2.** Comparison of general characteristics between GP and nurses.

|  | GP (n = 214) | Nurses (n = 526) | *P* value |
| --- | --- | --- | --- |
| **Gender, %** |  |  |  |
| Male | 86 (40.19) | 76 (14.45) | <0.001<sup>a</sup> |
| Female | 128 (59.81) | 450 (85.55) |  |
| **Age, median (IQR), yr** | 25 (22-38.3) | 29 (26-34) | <0.001<sup>b</sup> |
| **Hospital classification, %** | NA |  |  |
| Grade 3A |  | 387 (73.57) |  |
| Grade 3B |  | 66 (12.55) |  |
| Grade 2A |  | 53 (10.08) |  |
| Grade 2B |  | 9 (1.71) |  |
| Others |  | 11 (2.09) |  |
| **Years of working, median (IQR), yr** | NA | 8 (3-12) |  |
| **Departments, %** | NA |  |  |
| Internal medicine |  | 122 (23.19) |  |
| Surgery |  | 81 (15.4) |  |
| Emergency |  | 24 (4.56) |  |
| Critical care medicine |  | 128 (24.33) |  |
| Gynecology & Pediatrics |  | 131 (24.9) |  |
| Others |  | 40 (7.6) |  |
| **Professional titles, %** | NA |  |  |
| Nurse |  | 92 (17.49) |  |
| Senior nurse |  | 257 (48.86) |  |
| Nurse-in-charge |  | 148 (28.14) |  |
| Deputy chief or higher |  | 29 (5.51) |  |
| **Management work, %** | NA |  |  |
| Yes |  | 140 (26.62) |  |
| No |  | 386 (73.38) |  |
| **Education background, %** | NA |  |  |
| College degree |  | 82 (15.59) |  |
| Bachelor or higher degree |  | 444 (84.41) |  |
| **Marriage, %** |  |  | <0.001<sup>a</sup> |
| Unmarried | 124 (57.94) | 218 (41.44) |  |
| Married | 88 (41.12) | 299 (56.84) |  |
| Divorce or others | 2 (0.01) | 9 (1.71) |  |
| **Single-child, %** |  |  |  |
| Yes | 80 (37.38) | 221 (42.02) | 0.245<sup>a</sup> |
| No | 134 (62.62) | 305 (57.98) |  |
| **Fertility, %** |  |  | <0.001<sup>a</sup> |
| Yes | 84 (39.25) | 274 (52.09) |  |
| No | 130 (60.75) | 252 (47.91) |  |

Abbreviations: GP, general public; IQR, interquartile range; NA, not applicable.
<sup>a</sup> chi-square test; <sup>b</sup> Mann–Whitney U-test

**Table 3.** Comparison of vicarious traumatization severity between GP and nurses.

|  | GP (n = 214) | Nurses (n = 526) | Z scores | *P* value |
| --- | --- | --- | --- | --- |
| Vicarious traumatization | 75.5 (62-88.3) | 69 (56-85) | -2.396 | 0.017 |
| Physiological responses | 18 (13-24) | 18 (13-23) | -0.446 | 0.656 |
| Psychological responses | 57 (47-65.3) | 52 (42-62) | -3.002 | 0.003 |
| Behavioral responses | 15 (12-18) | 14 (11-17) | -2.764 | 0.006 |
| Emotional responses | 19 (15-23) | 17 (13-21) | -2.848 | 0.004 |
| Cognitive responses | 8 (6-10) | 8 (6-10) | -0.495 | 0.621 |
| Life beliefs | 13.5 (11-17) | 12 (10-15) | -2.877 | 0.004 |

Abbreviations: GP, general public.

### Comparison of general characteristics and VT scores between FLNs, nFLNs, and GP

In the study, a total of 526 nurses, which consist of 234 FLNs and 292 nFLNs, were enrolled. Results showed a statistical difference in hospital classification (*P* < 0.001), departments (*P* < 0.001), professional titles (*P* = 0.044), single child or not (*P* < 0.001), and fertile or not (*P* = 0.024) between FLNs and nFLNs (**Table 4**). Furthermore, the VT scores between FLNs and nFLNs were compared. The study found that VT scores [FLNs: 64 (IQR = 52–75), nFLNs: 75.5 (IQR = 63–92); *P* < 0.001] and their sub-items scores showed a significant increase in nFLNs than those of FLNs (**Table 5 and Figure 2**). The sub-item scores are as follows: physiological responses [FLNs: 17 (IQR = 12–21), nFLNs: 19 (IQR = 13.3–25); *P* < 0.001], psychological responses [FLNs: 46.5 (IQR = 38–55), nFLNs: 56.5 (IQR = 47–68.8); *P* < 0.001], behavioral responses [FLNs: 13 (IQR = 10–15), nFLNs: 15 (IQR = 12–18); *P* < 0.001], emotional responses [FLNs: 15 (IQR = 12–18.3), nFLNs: 19 (IQR = 15.3–23); *P* < 0.001], cognitive responses [FLNs: 7 (IQR = 5–9), nFLNs: 9 (IQR = 7–11); *P* < 0.001], and life beliefs [FLNs: 11 (IQR = 9–13), nFLNs: 14 (IQR = 11–17); *P* < 0.001]. Intriguingly, the VT scores between FLNs, GP, and nFLNs, were separately compared and showed significantly lower VT scores in FLNs than those in the GP and nFLN groups (*P* < 0.001). However, no significant difference was noted regarding VT scores between GP and nFLNs (*P* > 0.05).

**Table 4.** Comparison of general characteristics between FLNs and nFLNs.

|  | FLNs (n = 234) | nFLNs (n = 292) | *P* value |
| --- | --- | --- | --- |
| **Gender, %** |  |  | 0.147<sup>a</sup> |
| Male | 28 (11.97) | 48 (16.44) |  |
| Female | 206 (88.03) | 244 (83.56) |  |
| **Age, median (IQR), yr** | 29.5 (26-34) | 29 (25-34) | 0.353<sup>b</sup> |
| **Hospital classification, %** |  |  | <0.001<sup>a</sup> |
| Grade 3A | 152 (64.96) | 235 (80.48) |  |
| Grade 3B | 29 (12.39) | 37 (12.67) |  |
| Grade 2A | 38 (16.24) | 15 (5.14) |  |
| Grade 2B | 7 (2.99) | 2 (0.68) |  |
| Others | 8 (3.42) | 3 (1.03) |  |
| **Years of working, median (IQR), yr** | 8 (3.8-13) | 7.5 (3-11) | 0.187<sup>b</sup> |
| **Departments, %** |  |  | <0.001<sup>a</sup> |
| Internal medicine | 60 (25.64) | 62 (21.23) |  |
| Surgery | 39 (16.67) | 142 (48.63) |  |
| Emergency | 13 (5.56) | 11 (3.77) |  |
| Critical care medicine | 96 (41.03) | 32 (10.96) |  |
| Gynecology & Pediatrics | 12 (5.13) | 19 (6.51) |  |
| Others | 14 (5.98) | 26 (8.9) |  |
| **Professional titles, %** |  |  | 0.044<sup>a</sup> |
| Nurse | 31 (13.25) | 61 (20.89) |  |
| Senior nurse | 128 (54.7) | 129 (44.18) |  |
| Nurse-in-charge | 61 (26.07) | 87 (29.79) |  |
| Deputy chief or higher | 14 (5.98) | 15 (5.14) |  |
| **Management work, %** |  |  | 0.182<sup>a</sup> |
| Yes | 69 (29.49) | 71 (24.32) |  |
| No | 165 (70.51) | 221 (75.68) |  |
| **Education background, %** |  |  | 0.115<sup>a</sup> |
| College degree | 43 (18.38) | 39 (13.36) |  |
| Bachelor or higher degree | 191 (81.62) | 253 (86.65) |  |
| **Marriage, %** |  |  | 0.114<sup>a</sup> |
| Unmarried | 105 (44.87) | 113 (38.7) |  |
| Married | 123 (52.56) | 176 (60.27) |  |
| Divorce or others | 6 (2.56) | 3 (1.03) |  |
| **Single-child, %** |  |  | <0.001<sup>a</sup> |
| Yes | 78 (33.33) | 143 (48.97) |  |
| No | 156 (66.67) | 149 (51.03) |  |
| **Fertility, %** |  |  | 0.024<sup>a</sup> |
| Yes | 109 (46.58) | 165 (56.51) |  |
| No | 125 (53.42) | 127 (43.49) |  |

Abbreviations: FLNs, front-line nurses; nFLNs, non-front-line nurses.
<sup>a</sup> Chi-square test; <sup>b</sup> Mann–Whitney U-test

**Table 5.** Comparison of vicarious traumatization severity between FLNs and nFLNs.

|  | FLNs (n = 234) | nFLNs (n = 292) | Z scores | *P* value |
| --- | --- | --- | --- | --- |
| Vicarious traumatization | 64 (52-75) | 75.5 (63-92) | -7.156 | <0.001 |
| Physiological responses | 17 (12-21) | 19 (13.3-25) | -4.001 | <0.001 |
| Psychological responses | 46.5 (38-55) | 56.5 (47-68.8) | -7.785 | <0.001 |
| Behavioral responses | 13 (10-15) | 15 (12-18) | -5.578 | <0.001 |
| Emotional responses | 15 (12-18.3) | 19 (15.3-23) | -8.119 | <0.001 |
| Cognitive responses | 7 (5-9) | 9 (7-11) | -4.855 | <0.001 |
| Life beliefs | 11 (9-13) | 14 (11-17) | -8.529 | <0.001 |

Abbreviations: FLNs, front-line nurses; nFLNs, non-front-line nurses.

**Figure 2.**
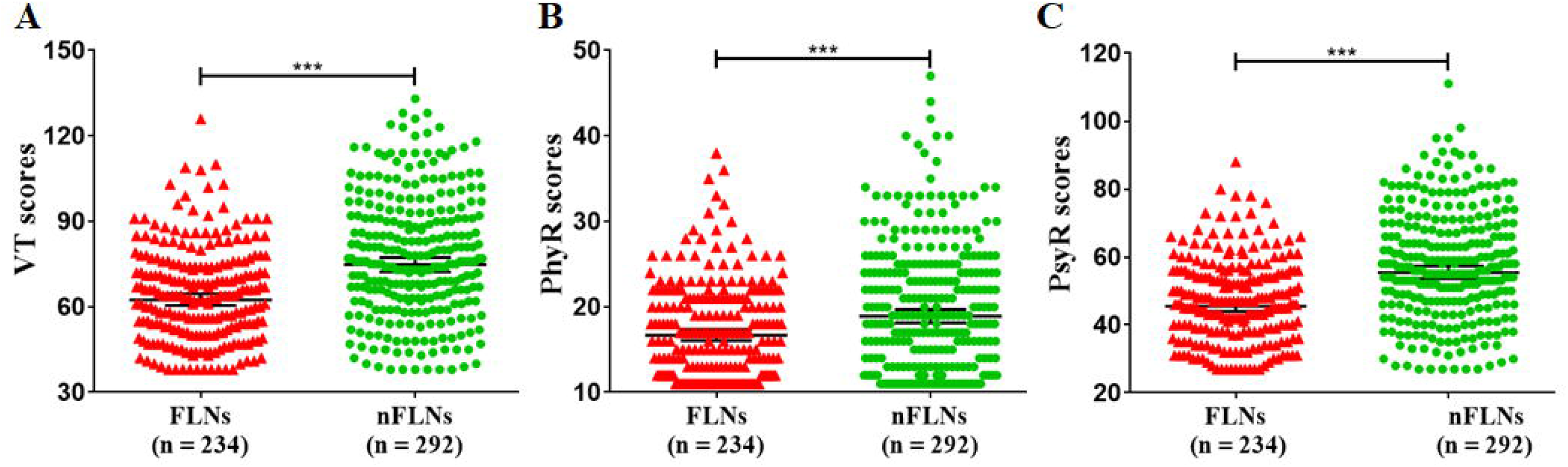
Scores for VT, PhyR, and PsyR between FLNs and nFLNs; VT scores (*P* < 0.001); PhyR scores (*P* < 0.001); PsyR (*P* < 0.001); FLNs, front-line nurses; nFLNs, non-front-line nurses; PhyR, physiological responses; PsyR, psychological responses; VT, vicarious traumatization

**Figure 3.**
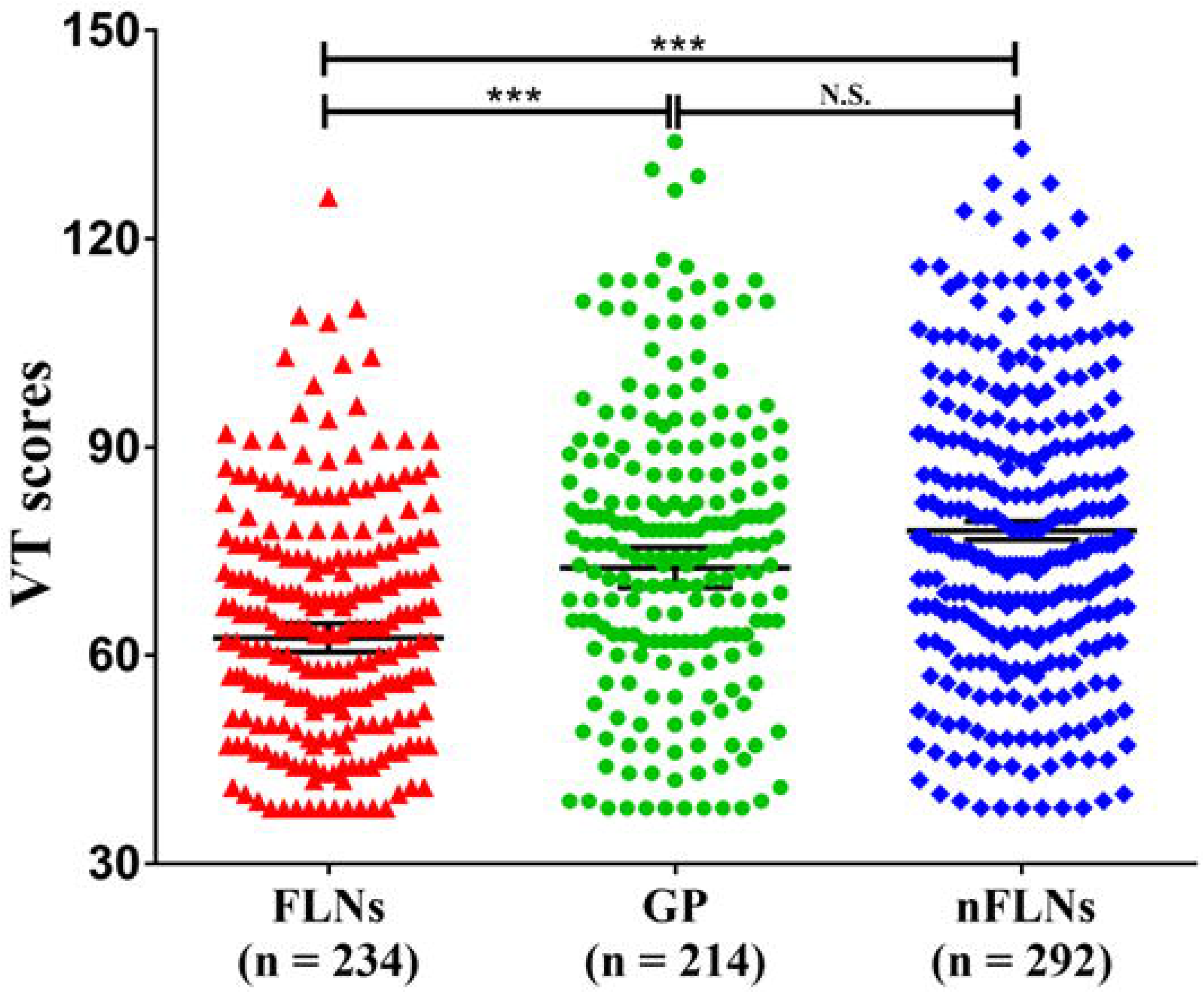
VT scores for FLNs, GP, and nFLNs; VT scores (*P* < 0.001). FLNs = front-line nurses; nFLNs, non-front-line nurses; VT, vicarious traumatization

### Risk factors for incidence of VT in nFLNs

Multiple linear regression analysis was performed to evaluate risk factors for the incidence of VT in nFLNs. Results showed that gender (OR = 3.117; 95% CI = 4.247–18.808; *P* = 0.002) and marriage status (OR = 2.072; 95%CI = 0.626–24.533; *P* = 0.039) are risk factors for VT of nFLNs (**Table 6**).

**Table 6.**
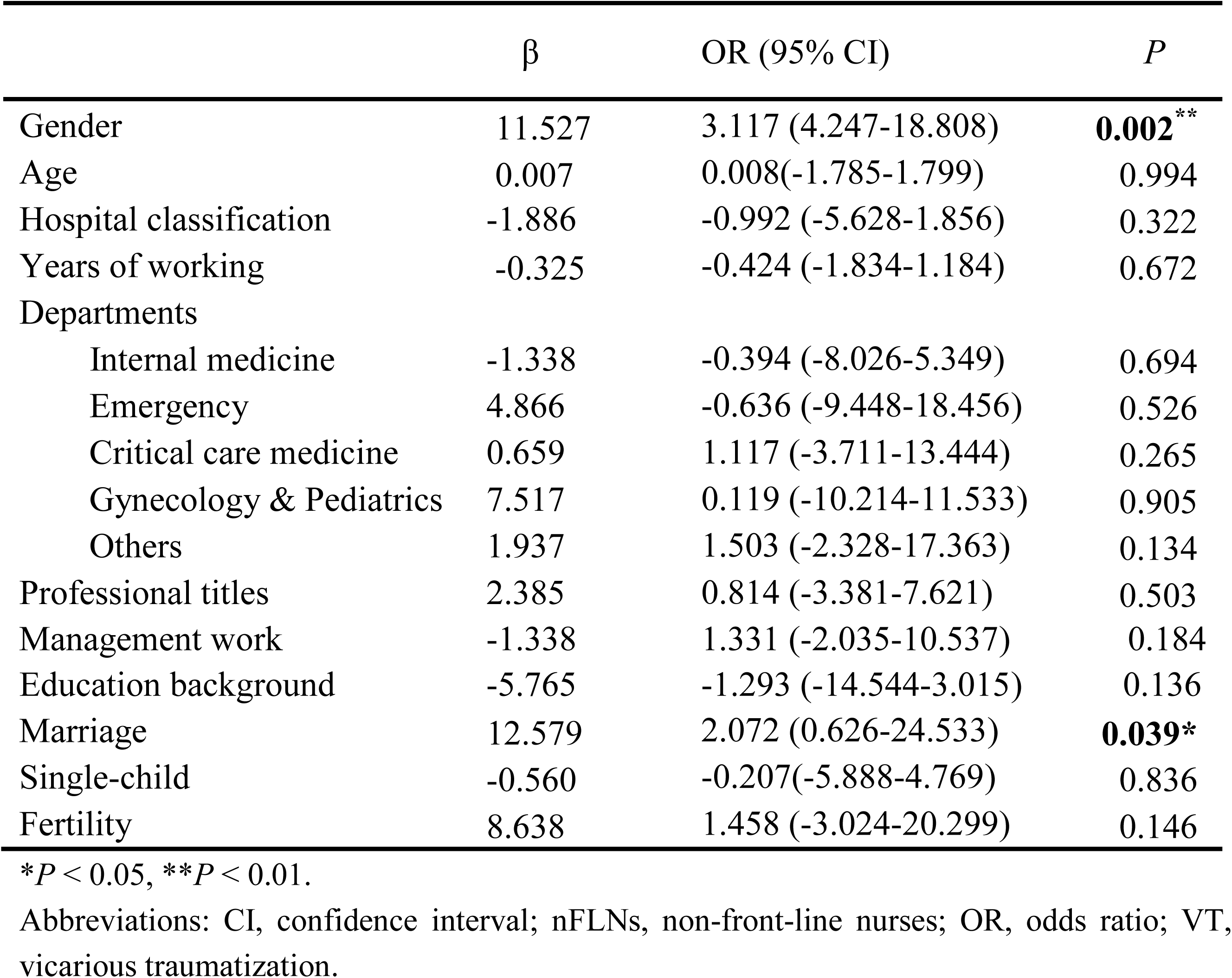
Risk factors for the incidence of VT in nFLNs.

## Discussion

The results showed that FLNs, nFLNs and the GP have experienced VT under the situation of the spread of COVID-19. However, the severity of VT in the three groups remains relatively differential. The study found that the level of VT in the GP was significantly higher than of nurses, including FLNs and nFLNs. Notably, although no significant differences were observed between the severity of VT in the nFLNs and the GP, its severity was significantly higher than that of the FLNs who came in close contact with patients with COVID-19. Therefore, the occurrence of psychological problems in front-line medical staff is likely, whereas the mental health of non-front-line medical staff and the GP should not be ignored. In addition, the study found that the severity of VT in nFLNs was more serious, whereas that of married and divorced or widowed nurses were higher than that of unmarried nurses. Most nurses are female, such that VT was more serious in female FLNs that that of male FLNs. These results suggest that female nurses with a marriage history are more likely to suffer from psychological disorders, which may be related to the fact that women are more sensitive to trauma^11,12^, and those with marriage history are largely older than those without marriage history. Interestingly, however, the study failed to find a correlation between age and increase in VT severity in nurses. To the best of the authors’ knowledge, this study is the first to focus on the psychological status, especially VT, in nurses aiding in COVID-19 control.

Although the severity of VT in the GP is higher than that of the medical staff, the study must emphasize that no difference was observed in the scores of physiological responses of VT between the two groups. A significant difference was noted for psychological responses, which consist of behavioral responses, emotional responses, and life beliefs. This finding may be highly related to the fact that China has adopted a strict isolation policy to deal with the epidemic, thus calling on the public to reduce face-to-face contact and communication to reduce the probability of viral transmission. During this period of COVID-19 proliferation, most of the GP stay at home for isolation. Thus, they gained more time to gather knowledge about the epidemic and the lives of other people, especially those of patients with COVID-19, through the Internet and media.^13^ The GP not only felt sympathy for patients with COVID-19, but are also excessively concerned about the medical staff. Importantly, the GP have no medical background and lack sufficient cognitive ability to increase awareness of the COVID-19 outbreak. In the issue of public health, psychological endurance is lacking. This notion suggests that during the spread and control of COVID-19, propaganda strategies should be well-organized and effective. In addition, early intervention measures should be taken to alleviate the psychological problems of the GP.^14^

The tendency of the severity of mental disorders to change over time has been widely reported.^15, 16^ Thus, the current study also verified this feature. FLNs with different working periods have different levels of VT severity. That is, the longer the support time, the more serious the alternative trauma. Although the difference fails to show statistical significance, it shows an increase trend. This finding may be highly related to the extension of working periods, where the medical staff consult with more patients with different processes of disease and varying degree of suffering. Under the long-term high-pressure and stress condition in fighting against the epidemic, their psychological defense will be negatively affected^17^, which can lead to increased VT severity. Therefore, adopting various levels of interventions for FLNs with VT is necessary during the diverse stages of medical support missions.

Results of the analysis indicate that the FLNs mainly originated from 3A and 3B hospitals (China’s hospital classification) and from critical care medicine and internal medicine departments. This group is mainly composed of middle-level backbone members, where most of them are single children and have not given birth. Close contact with patients with COVID-19 and direct exposure to the patients’ physical and psychological sufferings have been well recognized to render FLNs prone to suffer from VT; therefore, the society and psychotherapists should actively pay more attention to the psychological problems of FLNs.^18,19^ However, the results of the study imply that the VT severity of nFLNs, regardless of physical or psychological responses, is more serious than that of FLNs. This finding suggests that nFLNs are more likely to suffer from psychological problems, whereas the psychological endurance of FLNs is stronger. This notion may be due to the fact that FLNs are voluntarily selected and provided with sufficient psychological preparation. Second, the selected FLNs are mainly middle-level backbone staff with working experience and psychological capacity. In addition, the VT of FLNs is typically derived from sympathy for patients with COVID-19, whereas nFLNs not only feel sympathy for patients with COVID-19, but also bear the worry and sympathy for front-line colleagues. Finally, the FLNs are more knowledgeable about the epidemic than the GP and nFLNs. Therefore, a transparent announcement of epidemic information is very beneficial to social and psychological constructs and psychological intervention at a later time.^20^ Collectively, the abovementioned factors may be possible reasons for the higher severity of VT in nFLNs than in FLNs.

The study has certain limitations. First, the observational objects are mainly nurses. The reason for this option is that the proportion of nurses in the medical teams for COVID-19 control constitutes more than 70%, such that investigating nurses is representative. Secondly, this study is a descriptive cross-sectional one, which is unable to explore the causal linkage between factors. Therefore, carrying out a longitudinal large-sized intervention study and enrolling clinical doctors and other medical workers, such as technicians, is necessary to further explore the pathogenesis, therapeutic strategies, and mechanisms of VT.

In summary, the results suggest that the GP and medical staff suffer from VT. However, the VT of non-front-line medical staff is more serious than that of front-line medical staff. Therefore, early intervention of VT and psychological stress for the GP and medical staff, as well as the transparent announcement of epidemic information can facilitate the psychological treatment and control of COVID-19.

## Data Availability

We declared that all the original data are available.

## Acknowledgments

This work was supported by grants from the National Natural Science Foundation of China (grant numbers: 81703482 and 81974171 to C. Y.).

## Author contribution

*Concept and design:* Z. Li, J. Ge, M. Yang, J. Feng, R. Wang, C. Liu, C. Yang

*Acquisition, analysis, and interpretation of data:* Z. Li, J. Ge, M. Yang, J. Feng, M. Qiao, R. Jiang, J. Bi, G. Zhan, X. Xu, L. Wang, Q. Zhou, C. Zhou, Y. Pan, S. Liu, H. Zhang, J. Yang, B. Zhu, Y. Hu, K. Hashimoto, Y. Jia, H. Wang, R. Wang, C. Liu, C. Yang

*Drafting of the manuscript:* Z. Li, J. Ge, C. Yang

*Critical revision of the manuscript:* Z. Li, J. Ge, M. Yang, J. Feng, R. Wang, C. Liu, C. Yang

*Statistical analysis*: Z. Li, J. Ge, C. Yang

*Supervision:* R. Wang, C. Liu, C. Yang

